# Enhancing ADHD Prediction in Adolescents through Fitbit-Derived Wearable Data

**DOI:** 10.1101/2024.09.20.24314084

**Authors:** Muhammad Mahbubur Rahman

## Abstract

Attention-deficit/hyperactivity disorder (ADHD) is a prevalent neurodevelopmental condition characterized by a complex etiology. The diagnostic process for ADHD is often time-consuming and subjective in nature. Recent advancements in machine learning offer promising avenues for improving ADHD diagnosis automatically using various data sources. In this study, we harness Fitbit-derived physical activity measurements to investigate potential associations with ADHD and evaluate machine learning classifiers for their predictive accuracy in ADHD diagnosis. Our analysis involves a sample of 450 participants from the Adolescent Brain Cognitive Development (ABCD) study data release 5.0. We conduct correlation analyses to explore the connections between ADHD diagnosis and Fitbit-derived measurements, including sedentary time, resting heart rate, and energy expenditure. Subsequently, we employ multivariable logistic regression models to assess the predictive capability of these measurements for ADHD diagnosis. Furthermore, we train machine learning classifiers to achieve a diagnosis by automatically categorizing individuals into ADHD+ and ADHD-groups. Our correlation analysis unveils statistically significant associations between ADHD diagnosis and Fitbit-derived measurements, suggesting a potential link between physical activity patterns and ADHD. Importantly, multivariable logistic regression models demonstrate that some of the Fitbit measurements significantly predict ADHD diagnosis. Notably, our Random Forest machine learning classifier outperforms other classifiers with cross-validation accuracy (0.89), AUC (0.95), precision (0.88), recall (0.90), f1-score (0.89) and test accuracy (0.88), surpassing the performance of previous ADHD classification studies. These findings not only lay the groundwork for further exploration but also offer insights into the clinical integration of wearable data for a deeper understanding and improved identification of ADHD.

## 1. Introduction

Attention-deficit/hyperactivity disorder (ADHD) is a common neurodevelopmental disorder in childhood and may persist into adulthood, affecting about 9.8% of U.S. children^1^ and 4.4% of adults^2^. It often co-occurs with anxiety^1,3^ and depression^4^, highlighting its complexity.

Misdiagnosis can lead to substance abuse, lower education attainment, and legal issues^5–7^. However, diagnosing ADHD is hindered by barriers like limited understanding, time-consuming assessments, and subjectivity^8,9^. The co-occurrence of similar conditions adds to the challenge^10^. Machine learning approaches can leverage valuable evidential information in automatic ADHD diagnosis.

Many studies have applied machine learning to predict ADHD, using various data sources such as continuous performance test (CPT) variables^11^, pupillometric biomarkers and time series^12^, EEG measurements^13^, brain signals^14^, brain connectome topological information^15^, functional MRI^16^, symptom ratings and neuropsychological measures^17^, 3D MR images^18^, and fMRI from ABCD study^19^.

However, daily physical activity data from smartphones and wearables may be linked to ADHD. Some studies found increased physical activity levels in children with ADHD^20^, higher resting heart rate (RHR) and lower step count linked to increased internalizing symptoms^21^, associations between physical activity and mental health symptoms^22,23^, links between sedentary behavior and mental health^24^ and association between physical activity and executive function^25^.

A study based on self-reported data identified a link between ADHD symptoms and sedentary behavior^26^, while another study suggested that sedentary activities like reading and studying could enhance executive function and academic skills^27^. Research revealed higher heart rates in children with ADHD^28^ and elevated RHR in adults with ADHD on stimulant medication^29,30^. Studies indicated that stimulant medications reduced daily energy expenditure in children with ADHD^31^, individuals with ADHD had higher resting energy expenditure^32^, and higher energy expenditure in late adolescence was associated with lower ADHD symptom scores^33^.

In most machine learning studies for ADHD diagnosis, researchers primarily relied on either brain images or EHR collected in lab or hospital settings. This approach, while informative, poses several challenges, including high costs, time-intensive, and ethical concerns regarding the potential inclusion of sensitive personal information when training machine learning models. Additionally, many investigations exploring the relationship between various physical activities and ADHD faced limitations stemming from small sample sizes, potentially compromising the representativeness and generalizability of their findings. Moreover, a substantial portion of these studies relied on self-reported data, which introduced the risk of recall bias and inaccuracies, potentially failing to capture the full spectrum of sedentary time and energy expenditure, thereby impacting result precision. Furthermore, the majority of the data were collected from a single site, which could limit the broader applicability of the analysis.

Nonetheless, the collection of physical activity summaries could be significantly enhanced by leveraging smartphone sensors and wearable devices such as Fitbit and smart watch. Our study aimed to address these challenges by harnessing data from the Adolescent Brain Cognitive Development (ABCD) study, an extensive, long-term study encompassing 11,874 adolescents across 21 research sites in the United States. This dataset includes comprehensive Fitbit measurements, providing interesting daily and weekly physical activity summaries that cloud offer invaluable insights into sedentary time, RHR, and energy expenditure investigation for a significant number of adolescents, both with and without ADHD. The primary goal of our study was to investigate potential correlations between Fitbit measurements, including sedentary time, RHR, and energy expenditure, and ADHD diagnosis, as well as their predictive capabilities in ADHD diagnosis through the development of machine learning models.

Our primary contributions are as follows:

- Establishing associations between Fitbit measurements, including sedentary time, RHR, and energy expenditure, and ADHD diagnosis.
- Investigating the complex relationship between ADHD diagnosis and various independent Fitbit measurements.
- Developing predictive models for ADHD in adolescents using Fitbit measurements and conducting a comprehensive comparative analysis across multiple machine learning algorithms.
- Leveraging the extensive ABCD dataset to better understand ADHD diagnosis by analyzing daily and weekly physical activity summaries collected via wearable devices like Fitbit.

To the best of our knowledge, the integrated investigation of these Fitbit measurements from ABCD study to predict ADHD diagnosis represents a novel approach that has not been previously explored. Additionally, our modeling incorporates demographic information of adolescents, influenced by a study conducted by Nagata et al. within the ABCD framework, which demonstrated associations between sociodemographic variables and physical activities, such as step counts using Fitbit^34^. **Figure 1** shows the overview of our whole study.

**Fig. 1.**
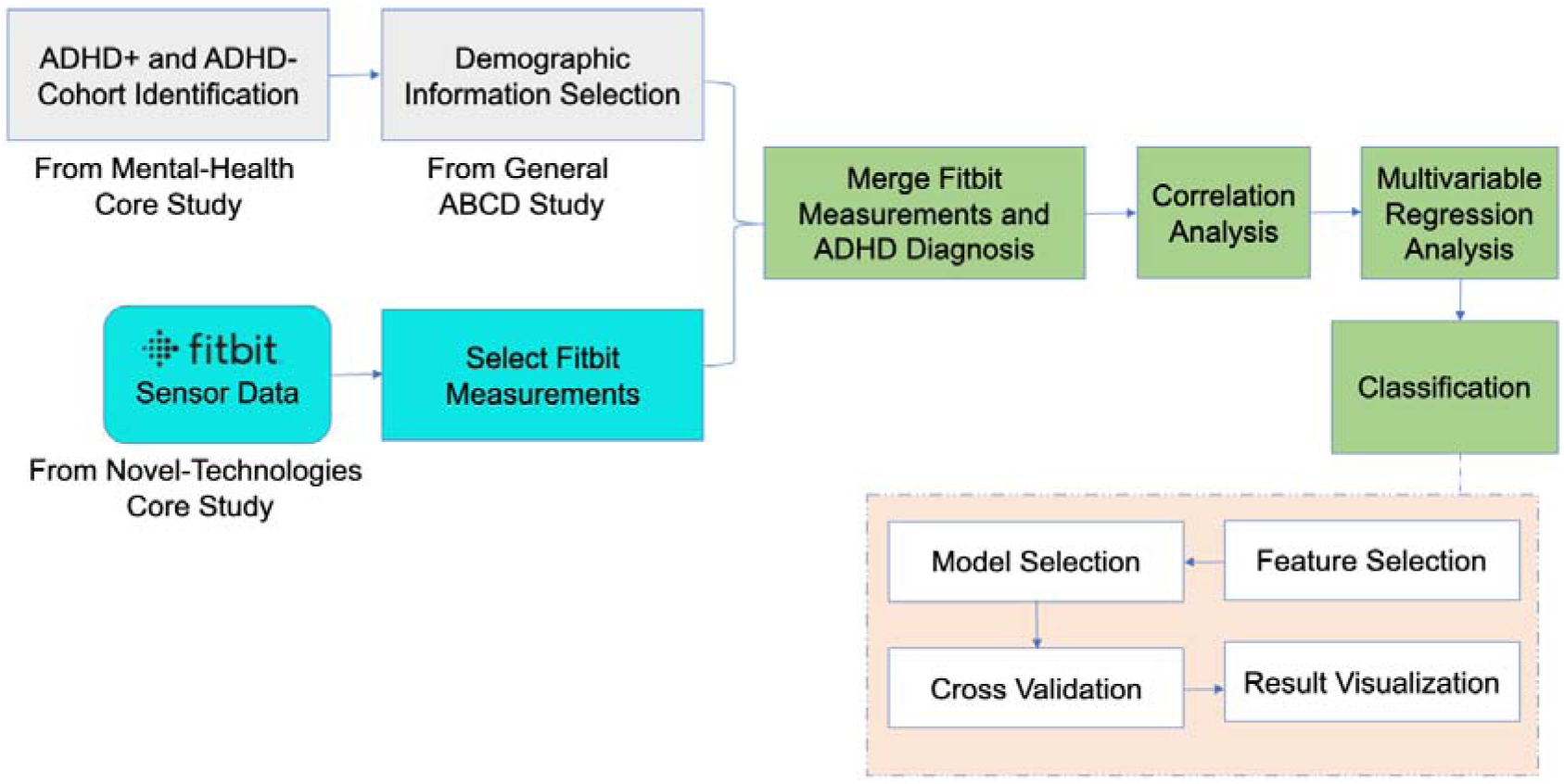
The overview of the whole study.

## 2. Materials and Methods

### 2.1. Study participants

In this study, the data were obtained from the ABCD research consortium. The ABCD study enrolled a total of 11,874 children, aged between 9 and 10, from 21 different study sites across the United States. For the purpose of our research, we used the ABCD Parent Diagnostic Interview for DSM-5 Full (KSADS-5) sub-study. In data release 5.0, the criteria for diagnosing ADHD were modified to necessitate impairment in two domains, as opposed to the earlier release that relied on impairment in only one domain. This sub-study enabled us to specifically identify subjects with ADHD positive (ADHD+) and ADHD negative (ADHD-) for our research cohort.

#### ADHD + group

Within the cohort of adolescents with ADHD, we included individuals who had a diagnosed ADHD condition at the time or who were in partial remission from ADHD. Additionally, we excluded any individuals who were diagnosed with ADHD in the past for a minimum of one school year but no longer exhibit ADHD symptoms (i.e., subjects who were fully in remission from ADHD). As detailed in the ABCD study’s data release 5.0, the determination of an ADHD diagnosis was calculated by evaluating impairment across at least two domains (e.g., the ability to engage in goal-directed behavior and the capacity to refrain from impulsive actions)^35^. A total of 357 individuals were identified as ADHD+ based on the inclusion and exclusion criteria.

#### ADHD-group

For the selection of ADHD-participants, we included adolescents who had never been diagnosed ADHD. Furthermore, we ensured that this group did not include individuals who were either partially or fully in remission from ADHD, nor those who were diagnosed with ADHD during any school year throughout their lifetime. However, we did not take into account the presence of any other mental health conditions when defining this ADHD-cohort. A total of 3311 unique individuals were identified as ADHD-based on the inclusion and exclusion criteria.

### 2.2. Fitbit measures

The ABCD Youth Fitbit daily physical activity summaries (n=7,439) involved the assessment of daily physical activity and sedentary behavior at the minute level, utilizing heart rate and accelerometer data from Fitbit sensors worn by adolescents. Additionally, the ABCD Youth Fitbit weekly physical activity summaries (n=7,076) captured weekly physical activity and sedentary behavior, including only days with adequate wear time for inclusion (>600 minutes of daytime wear) from the Fitbit sensors worn by adolescents. These datasets encompassed minutes spent in various activity intensities and recorded step counts, categorized into weekdays, weekends, daytime, nighttime, and all days of the week. Fitbit data was collected at baseline, the 2-year follow-up, and the 4-year follow-up using the Fitbit Charge 2 model worn on the wrist with parental consent. The participants wore Fitbit consistently for a period of over 21 days except during bathing and any water activities. Our study integrated Fitbit measurements across all three phases with minimal participants overlap. Data from both activity summaries were used in our Fitbit measurements of participant’s daily and weekly physical activity summaries, providing essential measurements relevant to our research goal. The specific measurements utilized in our study are shown in **Table 1**, fall within three primary categories: sedentary time, resting heart rate, and energy expenditure. The measurement definitions were directly taken from ABCD data.

**Table 1.**
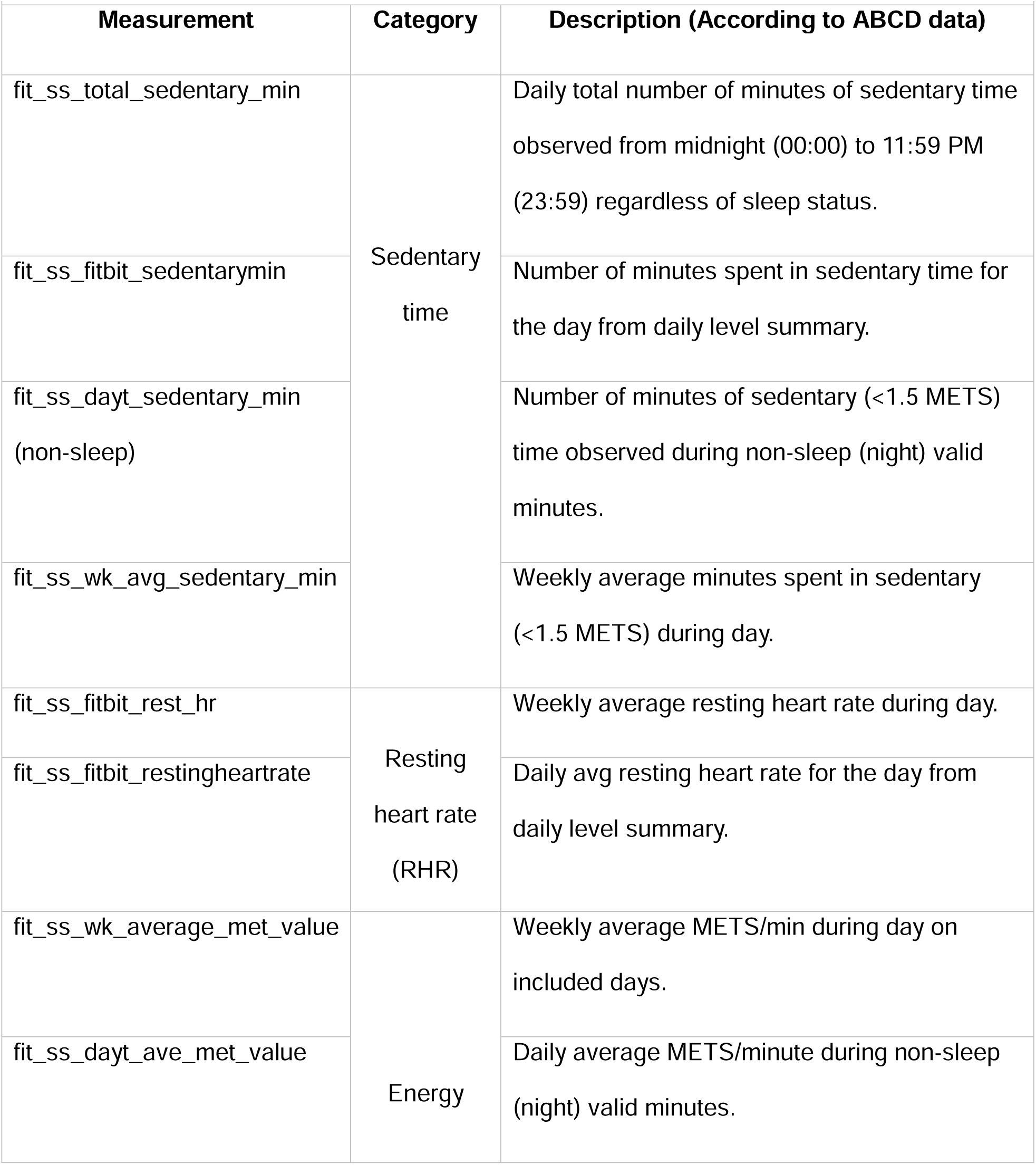
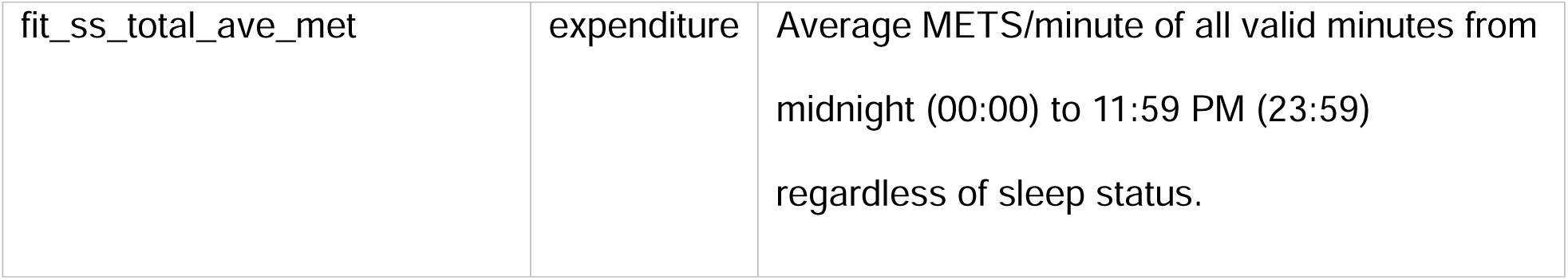
Fitbit measurements.

“Sedentary time” encompasses extended periods of inactivity or limited physical activity, signifying the duration spent in a seated, reclined, or lying position with minimal bodily movement and low energy expenditure. It commonly includes activities such as sitting at a desk, watching television, or using a mobile phone, where individuals engage in little to no physical effort. “Resting heart rate” represents the number of heart beats per minute during periods of rest. This measurement is typically taken while an individual is awake, in a state of relaxation, and not involved in any physical activity. Monitoring RHR serves as a useful tool for assessing overall well-being and tracking changes in fitness over time. “Energy expenditure,” expressed as METs/min (metabolic equivalents per minute), is a metric used to gauge the rate of energy consumption during various activities. An MET minute represents the energy expended within a minute of activity while at rest. To put it simply, 1 MET corresponds to the resting metabolic rate of an average adult, estimated at around 3.5 milliliters per kilogram per minute of oxygen consumption (ml/kg/min).

We conducted thorough data cleaning and preprocessing, eliminating rows with entirely missing values. If a single cell value was missing for a particular subject, we employed data imputation by replacing the missing data with the mean value of that specific subject’s variable. The detailed data processing steps are given in the **supplemental material: data processing and merging section**.

### 2.3. Study data sample

The Fitbit data was integrated with the ADHD+ samples, resulting in the identification of Fitbit data for specific measurements/variables in 225 out of the 352 ADHD+ subjects. Consequently, our final group of ADHD+ participants consisted of 225 distinct subjects. Likewise, among the 3,311 ADHD-subjects, 2,230 unique participants had corresponding Fitbit data, forming our ADHD-sample of 2,230 distinct subjects.

Given that our ADHD+ subject count reached 225, an effort was made to balance the participant list by selecting nearly equivalent ADHD-subjects from a pool of 2,230 candidates. This selection was performed through stratified sampling, considering factors like gender, age, race, parent’s education, marital status, and income level (**Supplemental material: Table 1**). This approach ensured a harmonized distribution between ADHD+ and ADHD-samples.

Ultimately, our dataset comprises 450 distinct adolescents, evenly split between ADHD+ and ADHD-subjects. The dataset is organized in a long format, featuring repeated observations for each subject over 21 days across different variables. It includes 10,045 Fitbit data records for 450 participants (ADHD+ and ADHD-), with a few having data beyond 21 days. Details on data merging, including explanations for retaining additional days for some participants, are available in the **supplemental material: data processing and merging section**.

### 2.4. Data analysis

In our study, statistical and predictive analyses were conducted. The statistical analysis was primarily carried out using correlation analysis. Additionally, confirmatory analysis, involving variable selection for machine learning models, was performed using multivariable logistic regression analysis. Subsequently, a predictive analysis was conducted using machine learning methods that included classification algorithms.

#### 2.4.1. Statistical analysis

Initially, a Pearson correlation analysis was conducted to examine the linear association between ADHD diagnosis (i.e., ADHD+ and ADHD-groups) and various Fitbit measurements. These Fitbit measurements included participants’ various energy expenditures, RHR, and sedentary time during different time intervals (**Table 1**). This correlation analysis was performed between-subjects comparisons, both with and without age, gender, race, parent’s education, and income level as control. A separate analysis using only age as a covariate was also conducted to observe its effects. Additionally, a repeated measures correlation analysis was conducted to explore within-subjects associations. To address potential false discoveries, a p-value correction using Holm’s Sequential Bonferroni method^36^ was applied. Corrections were applied to both between- and within-subjects analyses.

Subsequent to the initial correlation analysis, a multivariable logistic regression analysis was conducted using the Maximum Likelihood Estimation (MLE) method. This approach was chosen due to our focus on predicting binary outcomes at the participant level, where daily observations are considered independent. This analysis aimed to investigate the relationship between ADHD diagnosis and independent Fitbit measurements. However, in this analysis, the repeated measures were handled through the inclusion of time (date) as covariate in modeling. The results of this analysis informed the selection of variables for our machine learning model, designed to predict binary ADHD diagnosis outcomes. Additionally, we conducted mixed effects modeling to assess subject-specific variability. Collinearity was evaluated by calculating the Variance Inflation Factor (VIF) and managed separately by scaling the variables and employing Principal Component Analysis (PCA).

#### 2.4.2. Predictive analysis and classification

A range of supervised machine learning algorithms was implemented to predict ADHD diagnosis, with a focus on distinguishing between ADHD+ and ADHD-subjects. These algorithms included Decision Tree (DT), Random Forest (RF), Naïve Bayes (NB), AdaBoost (Ada) classifier, Light Gradient Boosting Machine (LGBM) classifier, Logistic Regression (LR) classifier, Support Vector Machines (SVM) classifier with non-linear kernels, and K Nearest Neighbors (KNN) classifier. To optimize the classification models, a grid search with 10-fold cross-validation was conducted, and a multi-core implementation was utilized to fine-tune the hyperparameters. Upon the identification of the best hyperparameters, the models were trained using these settings. To reduce the risk of overfitting in DT and RF, we applied diverse strategies such as pruning, optimizing the number of samples per leaf, and increasing the number of trees.

Our models were trained using Fitbit measurements that demonstrated statistical significance according to our multivariable logistic regression analysis, in addition to the demographic variables such as participants’ age, gender, race, parent’s socioeconomic status, and parent’s education. To prepare for training, essential preprocessing steps were taken, encompassing the removal of duplicates, conversion of categorical variables into numerical representations, and data normalization. These steps were essential to ensure compatibility with the classification algorithms.

The models were evaluated using a range of performance metrics. In the training process for all models, 10-fold cross-validation was applied. Furthermore, we partitioned the data into an 80:20 ratio, where the model was trained on 80% of the data, and its performance was evaluated on the remaining unseen 20% of the dataset. Accuracy, precision, recall, and F1-scores were computed using both the 10-fold cross-validation and training-test split procedures. Additionally, learning curves and ROC curves, along with AUC scores, were generated to assess the models’ ability to generalize.

Python 3.9 and R version 4.3.2 were utilized for all analyses. Data processing was carried out with the Pandas and Numpy libraries. Correlation analyses were performed using the Statsmodels libraries. Multivariable logistic regression and mixed effects regression analyses were conducted using lme4 and car packages. The development of machine learning models was done using the Scikit-Learn and LightGBM machine learning libraries, with multi-core processing to optimize efficiency. Data and results visualizations were created using the Matplotlib, Seaborn and Pandas packages.

## 3. Results

The study had 450 participants whose demographic details are presented in **Table 2**. **Supplementary Material: Table 3** provides demographic characteristics for the ADHD+ group, while **Supplementary Material: Table 4** presents corresponding information for the ADHD-(control) group.

**Table 2.**
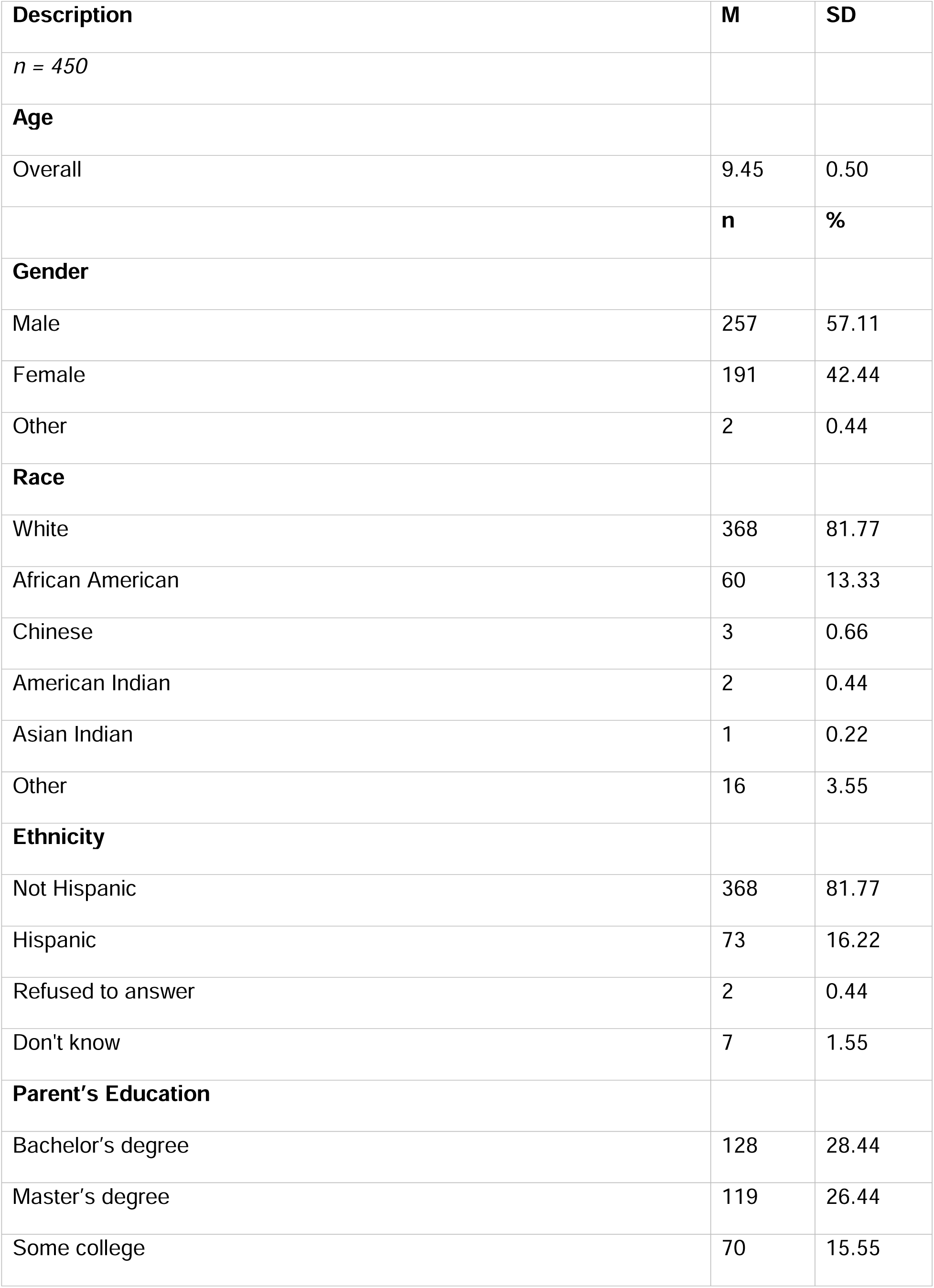
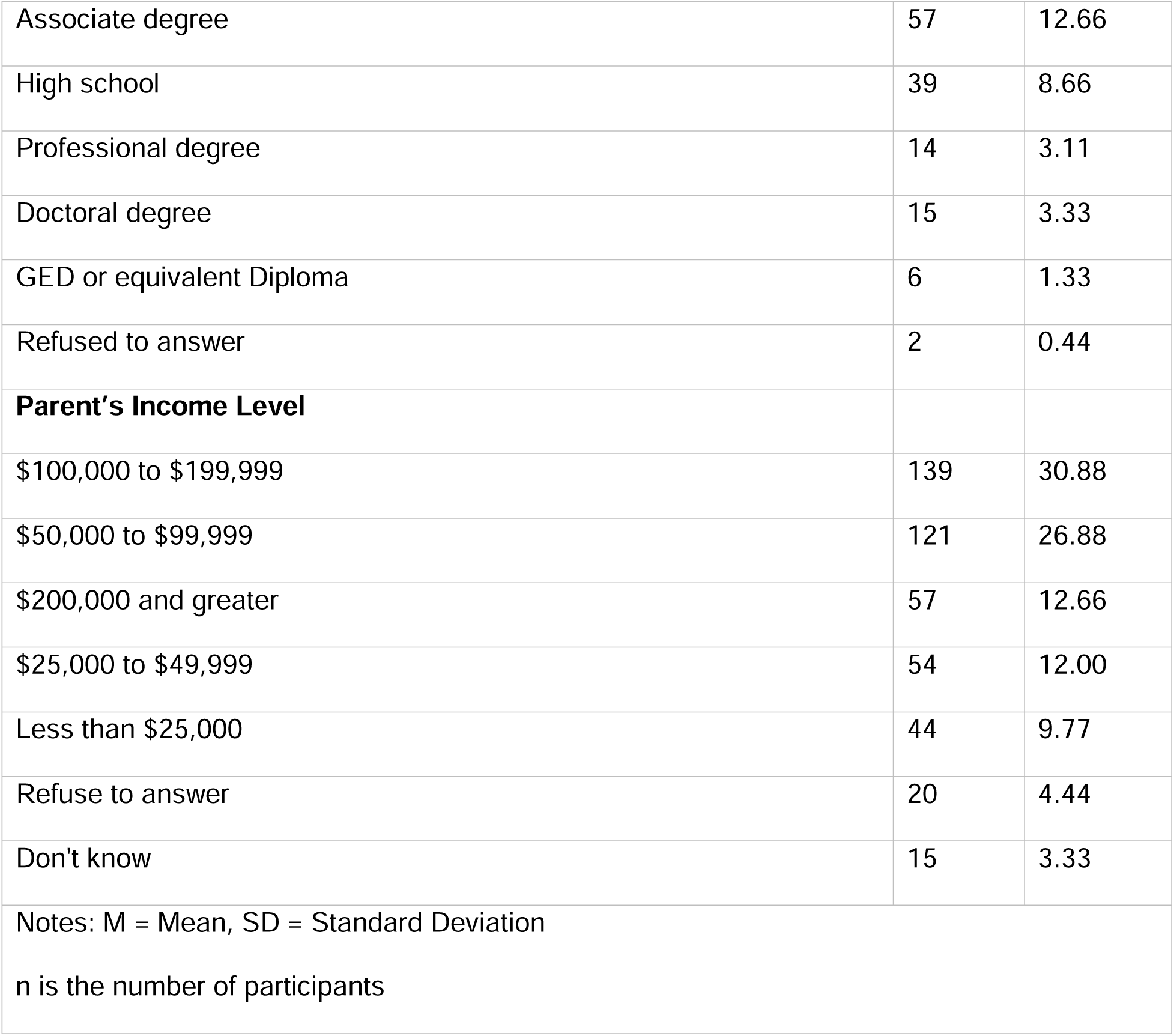
Participant’s Characteristics.

| **Description** | **M** | **SD** |
| --- | --- | --- |
| *n* = 450 |  |  |
| **Age** |  |  |
| Overall | 9.45 | 0.50 |
|  | **n** | **%** |
| **Gender** |  |  |
| Male | 257 | 57.11 |
| Female | 191 | 42.44 |
| Other | 2 | 0.44 |
| **Race** |  |  |
| White | 368 | 81.77 |
| African American | 60 | 13.33 |
| Chinese | 3 | 0.66 |
| American Indian | 2 | 0.44 |
| Asian Indian | 1 | 0.22 |
| Other | 16 | 3.55 |
| **Ethnicity** |  |  |
| Not Hispanic | 368 | 81.77 |
| Hispanic | 73 | 16.22 |
| Refused to answer | 2 | 0.44 |
| Don't know | 7 | 1.55 |
| **Parent's Education** |  |  |
| Bachelor's degree | 128 | 28.44 |
| Master's degree | 119 | 26.44 |
| Some college | 70 | 15.55 |
| Associate degree | 57 | 12.66 |
| High school | 39 | 8.66 |
| Professional degree | 14 | 3.11 |
| Doctoral degree | 15 | 3.33 |
| GED or equivalent Diploma | 6 | 1.33 |
| Refused to answer | 2 | 0.44 |
| **Parent's Income Level** |  |  |
| \$100,000 to \$199,999 | 139 | 30.88 |
| \$50,000 to \$99,999 | 121 | 26.88 |
| \$200,000 and greater | 57 | 12.66 |
| \$25,000 to \$49,999 | 54 | 12.00 |
| Less than \$25,000 | 44 | 9.77 |
| Refuse to answer | 20 | 4.44 |
| Don't know | 15 | 3.33 |
| Notes: M = Mean, SD = Standard Deviation |  |  |
| n is the number of participants |  |  |

**Table 3.** Correlation analysis.

| Fitbit measurements | Between participants |  |  |
| --- | --- | --- | --- |
|  | Without Controls | With Controls | Corrected p-value<br>(without Controls) |
| fit_ss_dayt_sedentary_min (non-sleep) | r: -0.041<br>p<0.0001 | r: -0.025<br>p = 0.013 | P<0.0001 |
| fit_ss_fitbit_sedentarymin | r: 0.070<br>p<0.0001 | r: 0.060<br>p<0.0001 | P<0.0001 |
| fit_ss_wk_avg_sedentary_min | r: -0.038<br>p<0.0001 | r: -0.016<br>p = 0.119 | P<0.0001 |
| fit_ss_total_sedentary_min | r: -0.081<br>p<0.0001 | r: -0.058<br>p<0.0001 | P<0.0001 |
| fit_ss_fitbit_rest_hr | r: 0.122<br>p<0.0001 | r: 0.161<br>p<0.0001 | P<0.0001 |
| fit_ss_fitbit_restingheartrate (day only) | r: 0.119<br>p<0.0001 | r: 0.160<br>p<0.0001 | P<0.0001 |
| fit_ss_dayt_ave_met_value | r: 0.041<br>p < 0.0001 | r: 0.003<br>p = 0.769 | P<0.0001 |
| fit_ss_wk_average_met_value | r: 0.058<br>p < 0.0001 | r: 0.004<br>p = 0.643 | P<0.0001 |
| fit_ss_total_ave_met | r: 0.056<br>p < 0.0001 | r: 0.019<br>p = 0.063 | P<0.0001 |

**Table 4.** Multivariable logistic regression model summary (with scaling).

| Variable | Coef | Std Err | Z-Score | P>\|Z\| |
| --- | --- | --- | --- | --- |
| Intercept | -0.05778 | 0.02028 | -2.850 | 0.004376 |
| Time | -0.10591 | 0.02155 | -4.914 | 8.94e-07 |
| fit_ss_total_sedentary_min | -0.13737 | 0.06533 | -2.103 | 0.035482 |
| fit_ss_fitbit_sedentarymin | 0.16524 | 0.04044 | 4.086 | 4.39e-05 |
| fit_ss_dayt_sedentary_min | 0.09526 | 0.05650 | 1.686 | 0.091804 |
| fit_ss_wk_avg_sedentary_min | 0.04500 | 0.03690 | 1.220 | 0.222554 |
| fit_ss_fitbit_rest_hr | 0.31254 | 0.10298 | 3.035 | 0.002406 |
| fit_ss_fitbit_restingheartrate | -0.05659 | 0.10298 | -0.549 | 0.582675 |
| fit_ss_wk_average_met_value | 0.14632 | 0.03873 | 3.779 | 0.000158 |
| fit_ss_dayt_ave_met_value | 0.23645 | 0.08517 | 2.776 | 0.005497 |
| fit_ss_total_ave_met | -0.18362 | 0.08236 | -2.230 | 0.025778 |

**Figure 2** shows measurements variability for weekly average sedentary time, RHR and energy expenditure with ADHD diagnosis. Based on the box plots and the presence of more data points above the maximum line in ADHD+ for both RHR and energy expenditure, there was indeed more variability in these Fitbit-derived measures among individuals with ADHD+ compared to ADHD-.

**Fig. 2.**
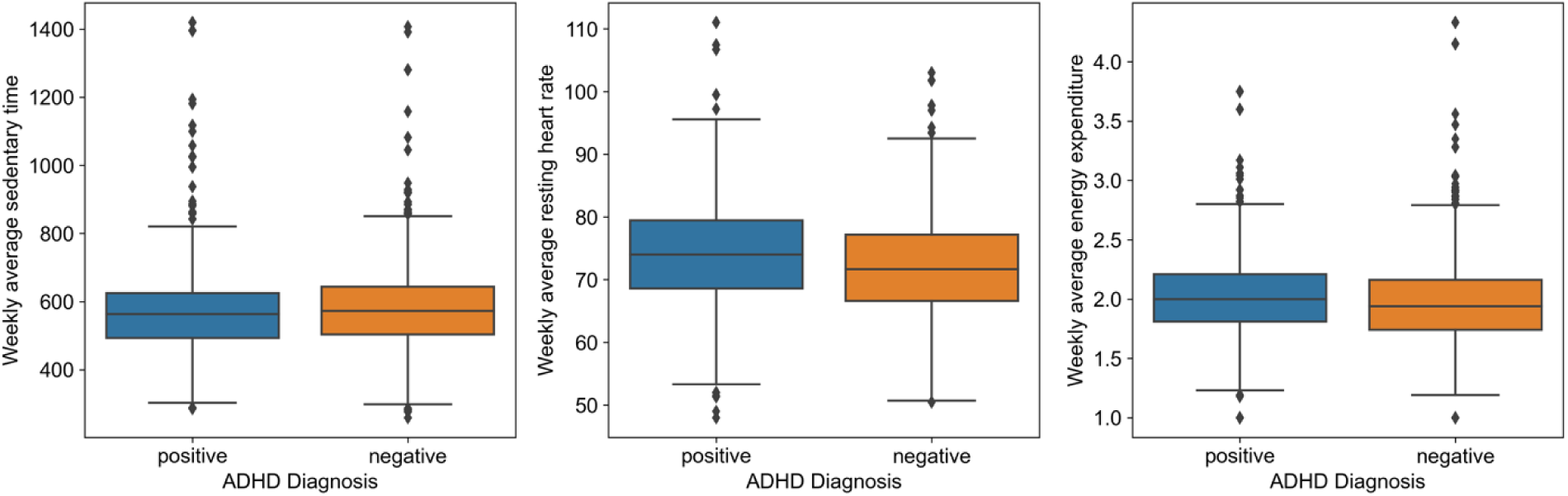
Fitbit measurements variability in ADHD diagnosis.

### 3.1. Association between diagnosis and measurements

During the Pearson correlation analysis conducted between ADHD diagnosis and Fitbit measurements without controls, statistically significant findings were obtained for between-participant analysis. However, the observed correlation coefficients were relatively small in magnitude. Statistically significant correlations were observed in most cases when demographics were used as controls. Notably, within-participant analyses demonstrated significance in specific scenarios shown in **supplemental material: Table 2**. Additionally, the results were also significance when age was employed as the sole covariate (**Supplemental material: Table 2**). **Table 3** provides the correlation analysis between ADHD diagnosis and Fitbit measurements.

#### 3.1.1. Sedentary time in minutes

The examination of sedentary time concerning ADHD diagnosis revealed interesting findings. Specifically, various aspects of sedentary time, including nighttime non-sleep (r = −0.041), daily total (r = −0.081), and weekly average (r = −0.038), demonstrated negative correlations with ADHD diagnosis. These results suggest that individuals with reduced overall sedentary time may exhibit a higher likelihood of an ADHD+ diagnosis. In contrast, daytime sedentary time exhibited a positive correlation (r = 0.070) with ADHD diagnosis. Importantly, these correlations retained significance even after controlling for demographics except for the weekly average.

However, when controlling for demographics, the coefficient values exhibited a slight decrease, suggesting that the control variables introduced a confounding influence on the observed relationship.

#### 3.1.2. Resting heart rate

The analysis of RHR in relation to ADHD diagnosis unveiled noteworthy outcomes. Notably, RHR measurements, both overall (r = 0.122) and during the day (r = 0.119), exhibited a substantial positive correlation with ADHD diagnosis. This implies that individuals with higher RHRs are more likely to receive an ADHD+ diagnosis. These correlations remained statistically significant even after controlling for demographics. The results were also significant in within-participants analyses (overall: r=0.032; during day: r=0.03). These findings emphasize the potential role of RHR as a significant marker associated with ADHD diagnosis.

#### 3.1.3. Energy expenditure

Higher energy expenditure while at rest, both for daily (r = 0.041) and weekly average (r = 0.058), along with the overall total energy expenditures (r = 0.056), displayed positive correlations with ADHD diagnosis. These correlations indicate that individuals with higher energy expenditures while at rest, are more likely to receive an ADHD+ diagnosis. These correlations were not statistically significant after controlling for demographics. However, the fact that controlling for these variables led to moderately lower coefficient values suggested that they exerted a noticeable influence on the relationship between ADHD diagnosis and energy expenditure.

### 3.2. Multivariable logistic regression modeling

In the multivariable logistic regression analysis, the predictors were Fitbit measurements and time (date) variable and the binary outcomes were ADHD diagnosis. The model’s results based on scaling the predictors are given in **Table 4**. We observed that six out of nine coefficient estimates were statistically significant, indicating the impact of these independent variables on the ADHD diagnosis. Despite the modest magnitudes of the coefficient values, our findings suggest a meaningful association between most of the Fitbit measurements and the ADHD diagnosis. The z-scores further support that these associations are unlikely to be the result of chance. We also conducted a multiple logistic regression model analysis with PCA for the overlapping variables (i.e., fit_ss_fitbit_rest_hr, fit_ss_fitbit_restingheartrate, fit_ss_dayt_ave_met_value, and fit_ss_total_ave_met). Two principal components out of these four variables demonstrated significance. The detailed results are shown in the **supplementary materials: Table 5**. Additionally, a mixed-effects model was trained; however, no significant results were obtained through mixed-effect regression analysis. The comprehensive results are provided in the **supplementary materials: >Table 6**.

**Table 5.**
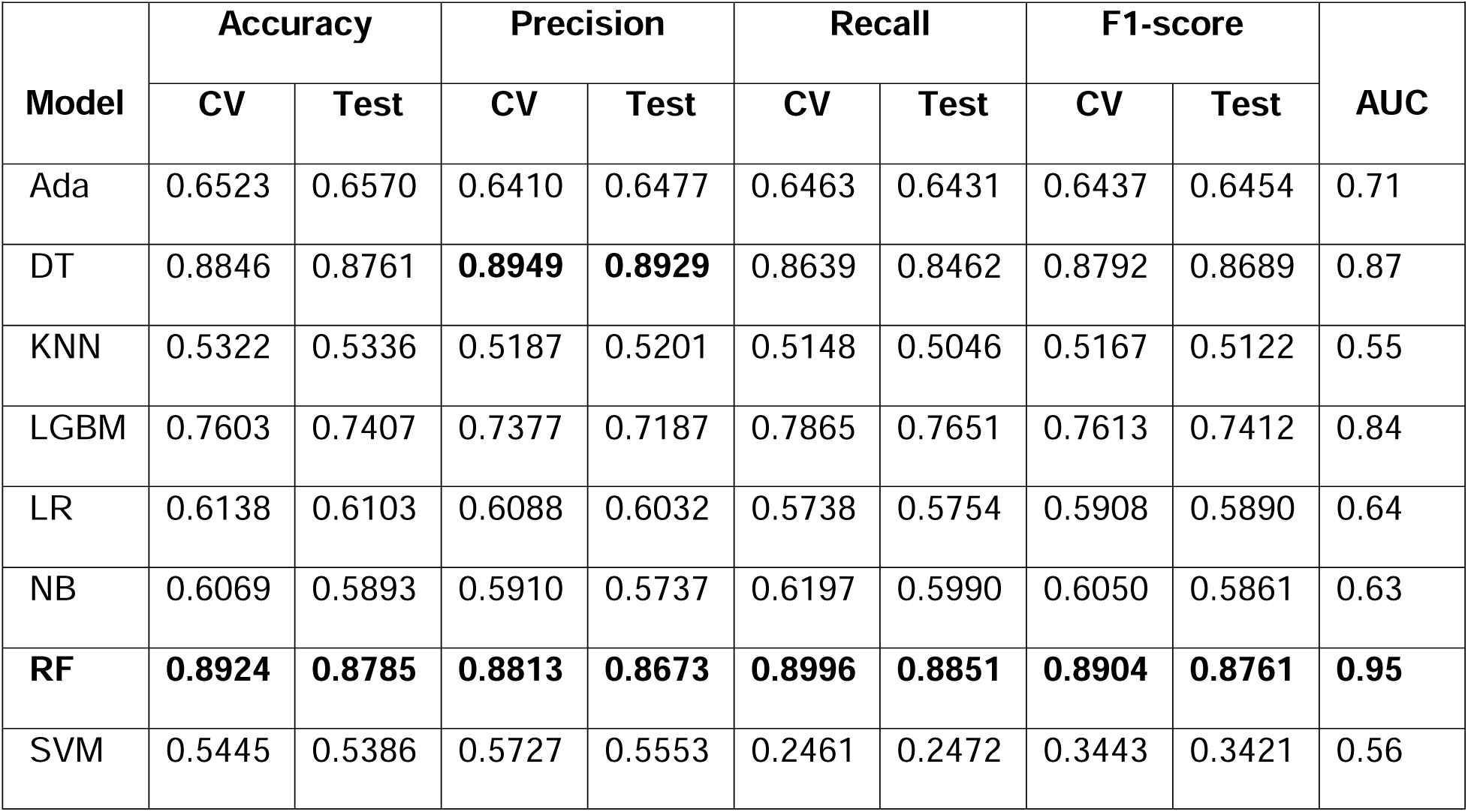
Classification Performance.

#### 3.2.1. Sedentary time in minutes

An increase in fit_ss_total_sedentary_min is associated with a decrease in the log-odds of ADHD diagnosis by 0.13737 units (z-score: −2.103, p = 0.0355). Conversely, an increase in fit_ss_fitbit_sedentarymin corresponds to a rise in log-odds by 0.16524 units (z-score: 4.086, p < 0.0001). Additionally, fit_ss_dayt_sedentary_min contributes to a log-odds increase of 0.09526 units, although this association is marginally significant (z-score: 1.686, p = 0.0918). Furthermore, fit_ss_wk_avg_sedentary_min does not show a significant association with ADHD diagnosis (z-score: 1.220, p = 0.2226).

#### 3.2.2. Resting heart rate

An increase in fit_ss_fitbit_rest_hr results in a rise of 0.31254 units in log-odds (z-score: 3.035, p = 0.0024). Conversely, an increase in fit_ss_fitbit_restingheartrate does not show a significant association with ADHD diagnosis (z-score: −0.549, p = 0.5827).

#### 3.2.3. Energy expenditure

An increase in fit_ss_wk_average_met_value corresponds to a log-odds rise of 0.14632 units (z-score: 3.779, p = 0.0002). Similarly, an increase in fit_ss_dayt_ave_met_value is associated with a log-odds rise of 0.23645 units (z-score: 2.776, p = 0.0055). Conversely, each unit increase in fit_ss_total_ave_met results in a decrease of 0.18362 units in log-odds (z-score: - 2.230, p = 0.0258).

The variables which had statistically significant associations in our multivariable logistic regression modeling between specific Fitbit measurements and ADHD diagnosis, were selected for the machine learning classification.

### 3.3. Classification and performance

**Table 5** summarizes machine learning classifier performance in predicting ADHD diagnosis using 10-fold CV and test dataset. RF outperformed other classifiers with 89.24% CV accuracy, 87.85% test accuracy, highest precision, recall, and F1-score, making it superior in ADHD diagnosis. In contrast, KNN underperformed with 53.22% CV accuracy and 53.36% test accuracy, indicating difficulty in distinguishing ADHD cases. Furthermore, strong AUC scores for RF ensemble methods indicated robust pattern learning from Fitbit data for ADHD prediction. **Figure 3** illustrates classifiers’ AUC scores.

**Fig. 3.**
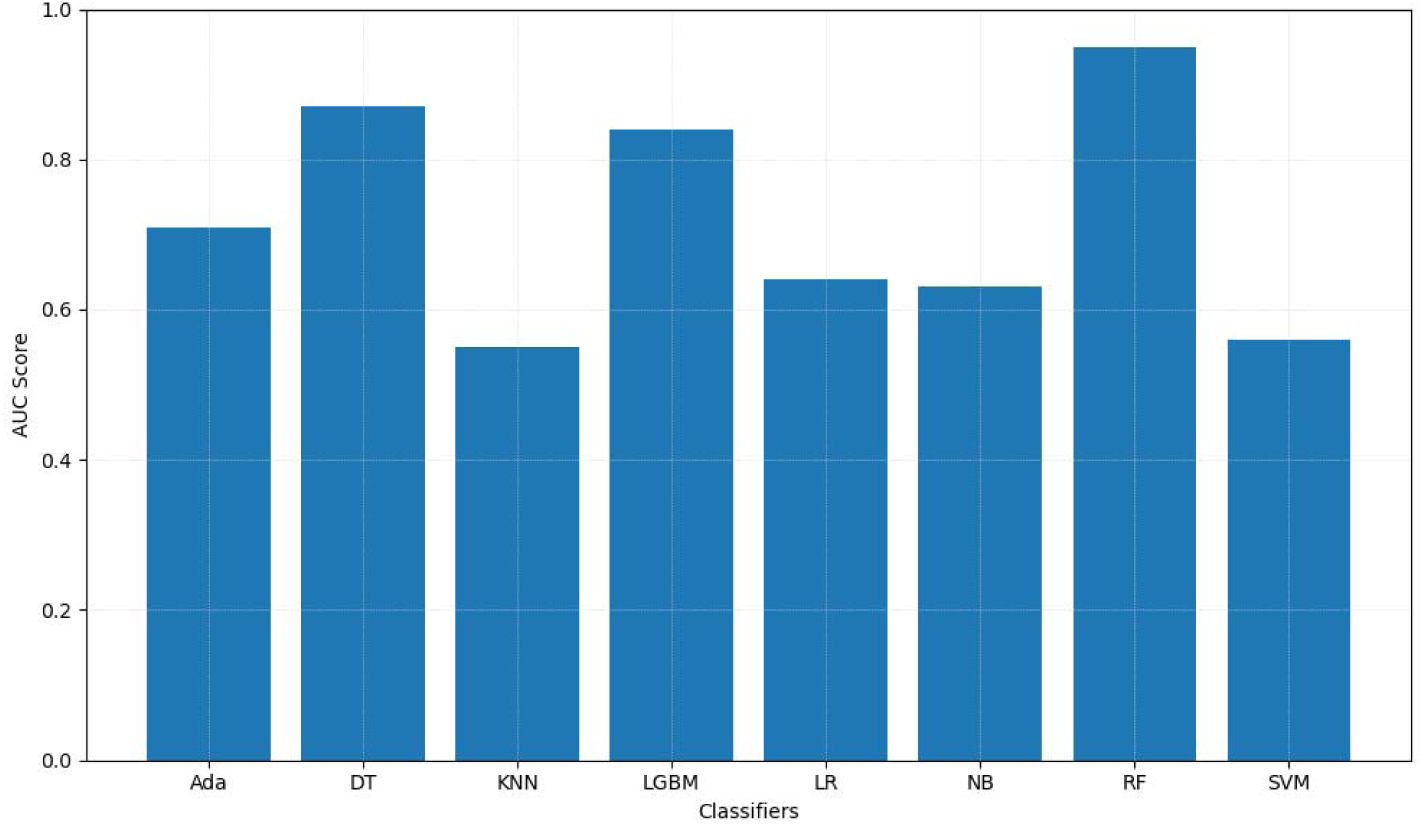
Classifiers’ AUC scores.

**Figure 4** depicts learning curves, illustrating training and validation performance across various classification algorithms. AdaBoost, while stable, displayed limited improvement, suggesting potential undergeneralization. In contrast, DT exhibited high initial accuracy, showcasing strong generalization and consistent improvement. KNN showed potential overfitting but reasonable generalization. LGBM learning curve initially overfitted, later generalized, stabilizing with balanced accuracy. LR demonstrated limited generalization, with minor accuracy gains. NB resembled LR in limited improvement. RF initially displayed overfitting, with subsequent improvement in validation accuracy, indicating a gradually improving generalization of the model over time. SVM displayed a pattern similar to that of LR. DT and RF are favored models for accurate ADHD prediction, while SVM may require further refinement to enhance performance.

**Fig. 4.**
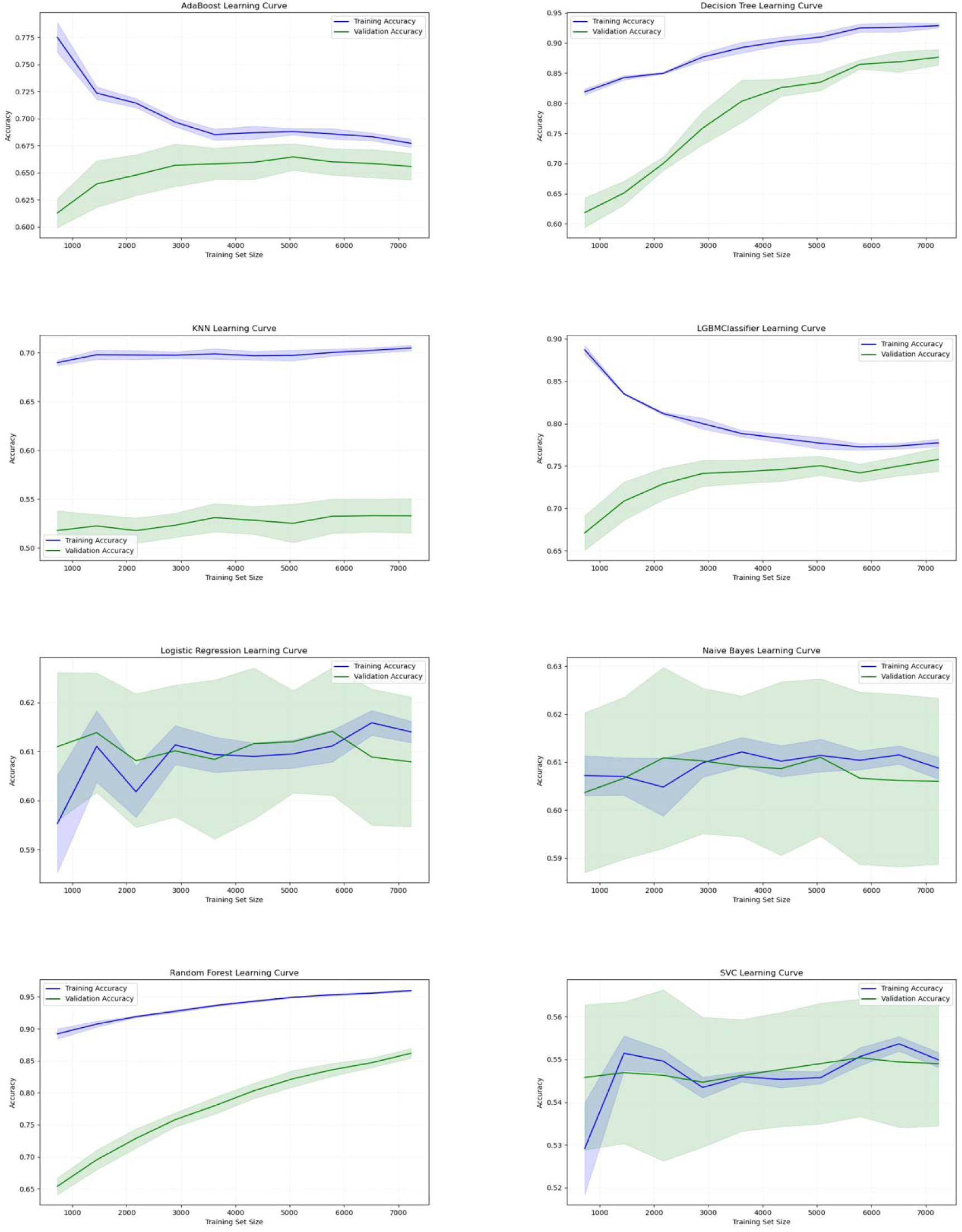
Learning curves for different classifiers.

In **Figure 5**, ROC curves illustrate the classifiers’ discriminative ability to distinguish ADHD+ and ADHD-cases. DT and LGBM stabilized gradually, whereas RF showed a rapid ascent and maintained high performance. In contrast, SVM, LR, and NB perform slightly better than random guessing. **Figure 6** indicates that RF, DT, and LGBM are the most promising models, showing stable performance with minimal variation across cross-validation folds.

**Fig. 5.**
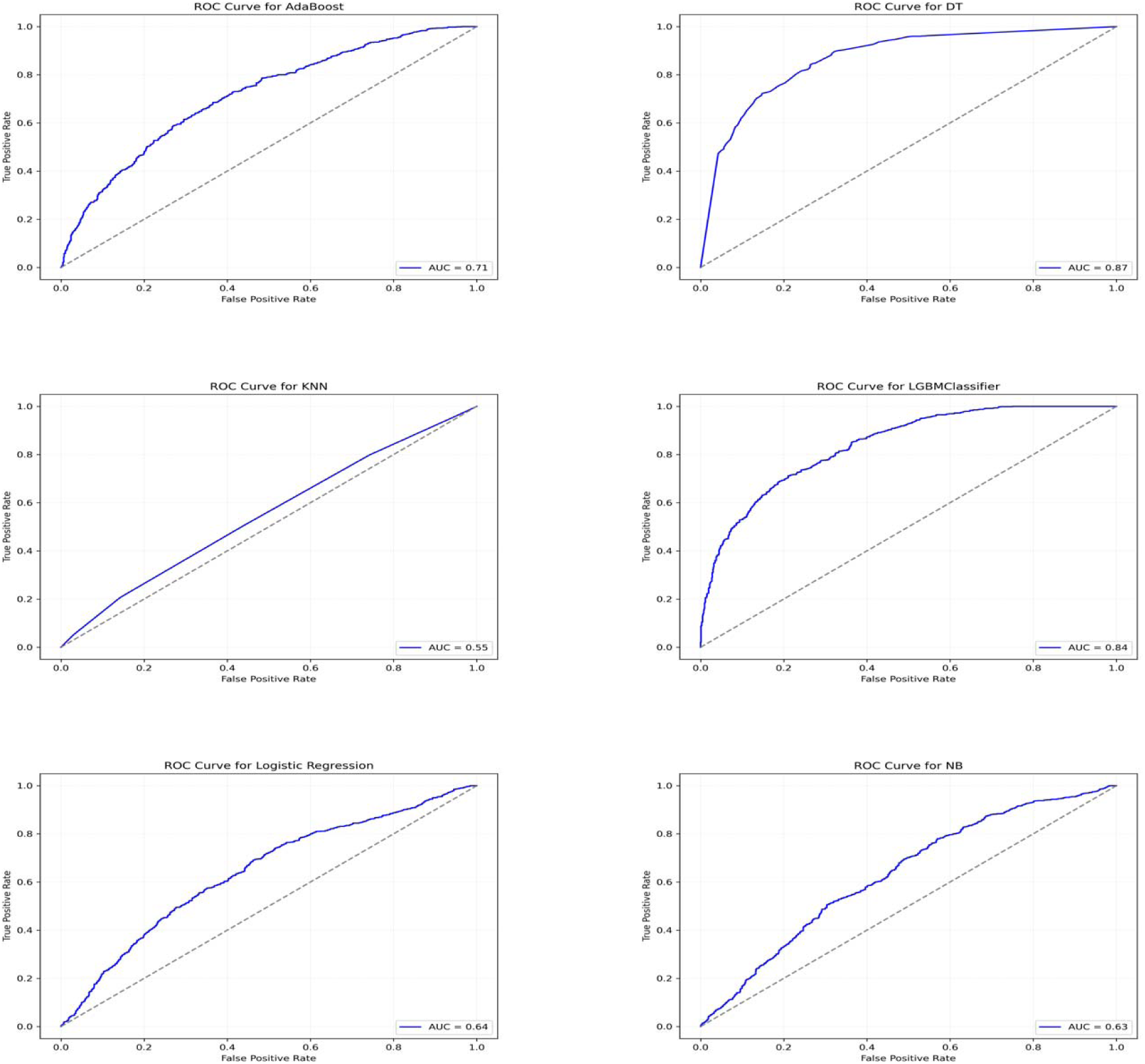
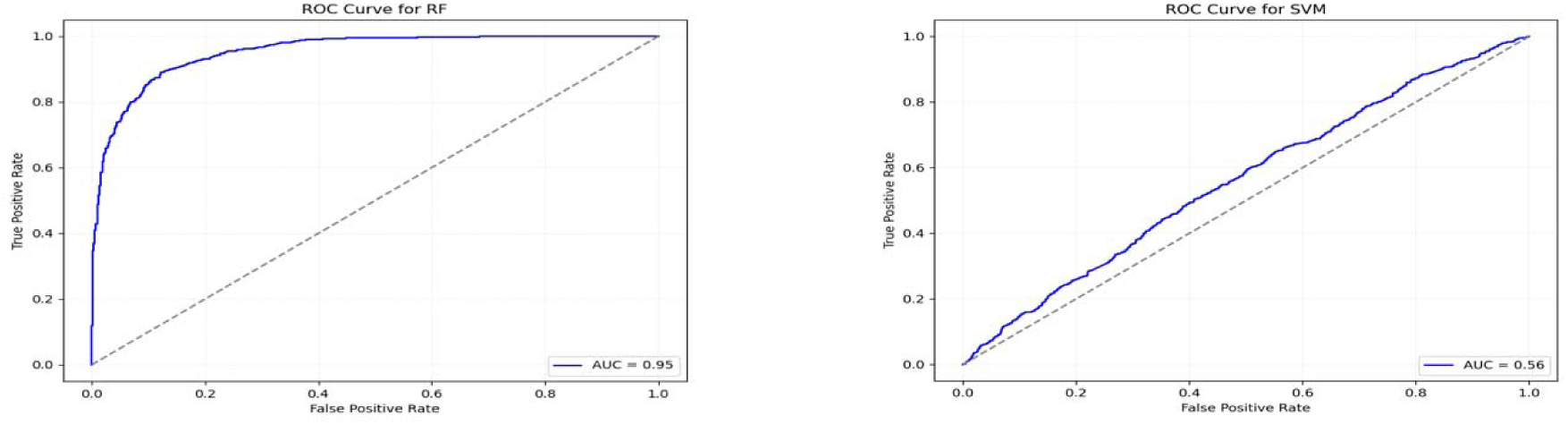
ROC curves for different classifiers.

**Fig. 6.**
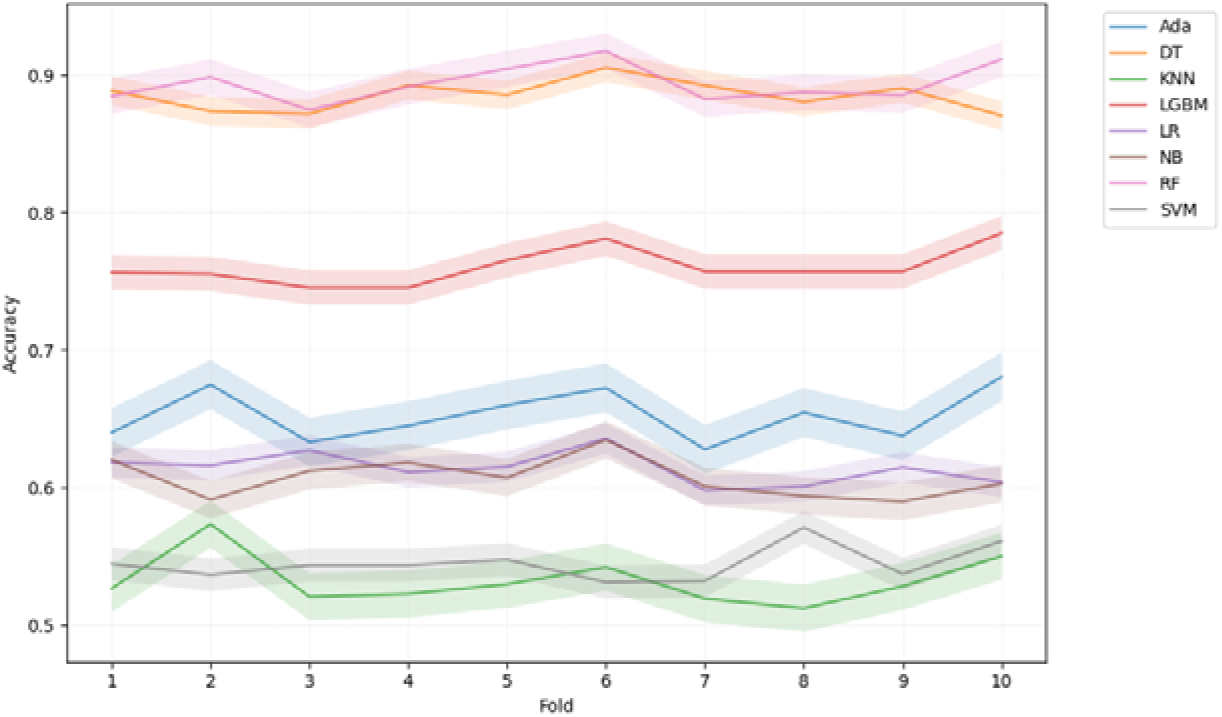
10-fold CV scores for different classifiers.

## 4. Discussion

The correlation analysis revealed statistically significant relationships between ADHD diagnosis and various Fitbit measurements, although the effect sizes were generally small in magnitude. These findings suggest that there is a statistical link between physical activity measures and ADHD, and the clinical significance of these associations should be explored. It is important to consider that the presence of mixed results in within-participants correlation analysis suggests variability in the relationships between variables across participants, potentially indicating heterogeneity, moderating factors, or complexities in the studied associations.

Regarding sedentary time, reduced overall sedentary time was associated with a higher likelihood of ADHD diagnosis, while increased daytime sedentary time showed a positive link to ADHD diagnosis. These findings highlight the importance of considering the timing of sedentary behavior when examining its relationship with ADHD. Our findings regarding increased daytime sedentary time align with previous research indicating a positive association between ADHD and sedentary behaviors^37–39^. In contrast, our results for overall sedentary time differ from the earlier studies, which may be attributed to differences in age groups and sample sizes.

However, it’s important to note that our study focused on the duration of sedentary time rather than specific sedentary activities. These disparities highlight the need for further investigation to gain a more comprehensive understanding of this pattern.

Our results indicated that individuals with higher RHR are more likely to have an ADHD. These results remained true even after controlling for demographics, suggesting that RHR could serve as a strong marker for ADHD diagnosis. Our findings are consistent with several prior studies that have reported a positive association between heart rate or RHR and ADHD in both children and adults when compared to those without ADHD^28–30^. Further research is needed to explore the underlying mechanisms of this relationship and its clinical implications.

Our analysis also revealed that individuals with higher energy expenditures while at rest may have an elevated likelihood of being diagnosed with ADHD. Importantly, these findings remained significant after controlling for demographics, with moderately lower coefficient values. Reduced coefficient values indicated that demographics had an impact on the association between ADHD and energy expenditure. These findings align with a prior study involving a different age group and smaller sample sizes^32^. However, they differ from another study^33^. These emphasize the necessity for further investigation to elucidate this relationship across different age groups, race, gender etc.

Additionally, the Fitbit measurements variability plots (**Figure 2**) provided interesting patterns, such as the greater variability in RHR and energy expenditure among ADHD+ individuals, suggested potential heterogeneity within this group.

The multivariable logistic regression analysis identified statistically significant associations between few specific Fitbit measurements, temporal factors, and binary ADHD diagnosis. Despite modest coefficients, sedentary time, resting heart rate, and energy expenditure emerged as influential factors. Notably, sedentary time showed nuanced associations with ADHD diagnosis. The temporal variable (time/date) played a significant role. Principal component analysis highlighted the importance of certain overlapping variables in predicting ADHD outcomes. Although the mixed-effects model did not show significant results, the findings emphasized the potential utility of Fitbit measurements and temporal considerations in understanding and predicting ADHD diagnoses.

Among all the classifiers trained in our study, RF consistently outperformed the other classifiers across accuracy, precision, recall, F1-score, and AUC. These results underscore the robustness and dependability of the RF model in accurately distinguishing between the ADHD+ and ADHD-groups. Notably, our top-performing classifier surpassed the performance of a previous study that used a similar sample from the ABCD study, integrating multiple measures of Resting-State Functional Magnetic Resonance Imaging (rsfMRI) in adolescent brains, achieving an accuracy of 0.6916 and an AUC of 0.7408^19^. Furthermore, our classifier outperformed another study conducted on a separate sample of 240 children, which utilized the temporal variability of dynamic functional connectivity from MRI brain images, achieving an accuracy of 0.78 and an AUC score of 0.84^40^. Both studies relied on expensive brain imaging methods and lab setups. In another study, Slobodin et al. achieved an accuracy of 0.87 using CPT data based on 458 children^11^. However, even their best classifier fell short of our top-performing classifier in terms of accuracy, precision, and recall scores. Our classifier also outperformed an SVM classifier trained by Das et al.^12^, which achieved an accuracy of 0.762 and an AUC score of 0.85 using pupillometric biomarkers and time series data.

In our analysis, KNN demonstrated the weakest performance. This suggests that KNN may not have effectively generalized the dataset, leading to difficulties in distinguishing between ADHD+ and ADHD-cases. We also observed high AUC scores for LGBM, DT and RF ensemble methods, indicating their effective learning of underlying data patterns. This highlights their suitability for predicting ADHD based on participants’ daily and weekly physical activity summaries collected through Fitbit.

Based on learning curve analysis, it could be inferred that DT and RF models demonstrated strong generalization, while KNN and SVM exhibited limitations in capturing complex data patterns. ROC curve analysis further confirmed the discriminative power of these classifiers, with DT, RF, and LGBM achieving high AUC scores, indicating their proficiency in distinguishing between ADHD+ and ADHD-cases. On the other hand, KNN had a lower AUC score, suggesting its challenges in effectively classifying the two groups.

Overall, our machine learning experiments highlight the effectiveness of ensemble methods, particularly RF, in accurately predicting ADHD diagnosis using participants physical activity summaries collected through Fitbit. These findings provide valuable insights into the choice of classifier for ADHD classification using Fitbit measurements, such as sedentary time, RHR and energy expenditure while in rest and suggest avenues for further research and model refinement in this clinical context.

## 5. Limitation and future work

This study is based on data obtained from the ABCD study, which may not capture the entire spectrum of children and adolescents across various age groups. We concentrated on binary classification of ADHD diagnosis (e.g., ADHD+ and ADHD-groups), without delving into potential ADHD subtypes. The current analysis employs the ABCD study’s ADHD diagnosis definition based on impairment in two domains, deviating from DSM criteria. In future investigations, we aim to broaden our scope by considering other conditions alongside the ABCD ADHD diagnosis definition to establish a DSM match. Additionally, our use of Fitbit measures in this study was confined to pre-existing data available in the ABCD dataset. Future research endeavors seek to expand our exploration by incorporating raw Fitbit data, uncovering additional variables related to ADHD treatment, sedentary time, resting heart rate (RHR), and energy expenditures. We also aim to deepen our understanding of ADHD, potentially leveraging advanced techniques such as deep learning.

## 6. Conclusion

This study illuminates the associations between Fitbit-derived physical activity summaries and ADHD diagnosis (e.g., ADHD+ and ADHD-groups) using the ABCD dataset. Our findings demonstrate that wearable technology, showed by the performance of the Random Forest classifier, holds promise in the realm of ADHD prediction and diagnostic applications. Our results provide a foundation for further exploration and the eventual integration of wearable data into the clinical landscape, fostering a deeper understanding of ADHD and advancing the accuracy of its identification.

## Conflict of Interest

The author declares no competing interests.

## Data Availability

All data produced are available online at https://data-dict.abcdstudy.org/?

## Acknowledgements

Data used in the preparation of this article were obtained from the Adolescent Brain Cognitive DevelopmentSM (ABCD) Study (https://abcdstudy.org), held in the NIMH Data Archive (NDA). This is a multisite, longitudinal study designed to recruit more than 10,000 children age 9-10 and follow them over 10 years into early adulthood. The ABCD Study® is supported by the National Institutes of Health and additional federal partners under award numbers U01DA041048, U01DA050989, U01DA051016, U01DA041022, U01DA051018, U01DA051037, U01DA050987, U01DA041174, U01DA041106, U01DA041117, U01DA041028, U01DA041134, U01DA050988, U01DA051039, U01DA041156, U01DA041025, U01DA041120, U01DA051038, U01DA041148, U01DA041093, U01DA041089, U24DA041123, U24DA041147. A full list of supporters is available at https://abcdstudy.org/federal-partners.html. A listing of participating sites and a complete listing of the study investigators can be found at https://abcdstudy.org/consortium_members/. ABCD consortium investigators designed and implemented the study and/or provided data but did not necessarily participate in the analysis or writing of this report. This manuscript reflects the views of the authors and may not reflect the opinions or views of the NIH or ABCD consortium investigators.

## Notes

### Competing Interest Statement

The authors have declared no competing interest.

### Funding Statement

This study did not receive any funding.

### Author Declarations

Institutional Review Board (IRB) of Children's National Hospital waived ethical approval for this work.

