## Supplemental for "Enhancing ADHD Prediction in Adolescents through Fitbit-Derived Wearable Data"

Supplementary Materials:

| **Table 1.** Demographic information and mapping | | |
| --- | --- | --- |
| **Original variable** | **Mapped to** | **Options** |
| demo_gender_id_v2 | Gender | 1 = Male, 2 = Female, 3 = Trans male, 4 = Trans female, 5 = Gender queer, 6 = Different, 777 = Refuse to answer, 999 = Don't know |
| demo_brthdat_v2 | age | Number in year |
| demo_race_a_p___10:  demo_race_a_p___11:  demo_race_a_p___12:  demo_race_a_p___13:  demo_race_a_p___14:  demo_race_a_p___15:  demo_race_a_p___15:  demo_race_a_p___17:  demo_race_a_p___18:  demo_race_a_p___19:  demo_race_a_p___20:  demo_race_a_p___21:  demo_race_a_p___22:  demo_race_a_p___23:  demo_race_a_p___24:  demo_race_a_p___25: | race | White; 0 = No; 1 = Yes  African American; 0 = No; 1 = Yes  American Indian; 0 = No; 1 = Yes  Asian Indian; 0 = No; 1 = Yes  Chinese; 0 = No; 1 = Yes  Other Asian: 0 = No; 1 = Yes  Other race; 0 = No; 1 = Yes  **Mapping race**: White, African American, American Indian, Asian Indian, Chinese, Other |
| demo_prnt_ed_v2 | education | 0: "Never attended", 1:"High school", 2:"High school", 3:"High school", 4:"High school", 5:"High school", 6:"High school", 7:"High school", 8:"High school", 9:"High school", 10:"High school", 11:"High school", 12:"High school", 13:"High school", 14:"GED or equivalent Diploma", 15:"Some college", 16:"Associate degree", 17:"Associate degree", 18:" Bachelor degree", 19: "Master degree", 20: "Professional degree", 21:"Doctoral degree", 777:"Refused to answer" |
| demo_comb_income_v2 | Income | 1: "Less than $25,000", 2: "Less than $25,000", 3: "Less than $25,000",4: "Less than $25,000", 5: "$25,000 to $49,999", 6:"$25,000 to $49,999",7:"$50,000 to $99,999", 8:"$50,000 to $99,999", 9:"$100,000 to $199,999", 10:"$200,000 and greater", 999:"Don't know", 777:"Refuse to answer" |
| demo_prnt_marital_v2 | Marital status | 1:"Married", 2:"Widowed", 3:"Divorced", 4: "Separated", 5: "Never married", 6:"Living with partner", 777:" Refused to answer" |
| demo_gender_id_v2 | Gender | 1: "Male", 2:"Female", 3:"Other", 4:"Other", 5:"Other", 6:"Other", 777:"Other", 999:"Other" |
| demo_ethn_v2 | Ethnicity | Hispanic; 1 = Yes; 2 = No ; 777 = Refuse to answer; 999 = Don't know |

| **Table 2.** Correlation analysis between ADHD diagnosis and Fitbit measurements for within-participants (with p-value correction) and with age as covariate | | | |
| --- | --- | --- | --- |
| **Fitbit Measurements** | **r (Within participants)** | **Corrected p-value (within participants)** | **Bewteen participants (with age as covariate)** |
| fit_ss_wk_average_met_value | -0.039  P = 0.0001 | 0001 | 0.059  P < 0.0001 |
| fit_ss_fitbit_rest_hr | 0.032  P = 0.0015 | 0.002 | 0.122  P < 0.0001 |
| fit_ss_wk_avg_sedentary_min | 0.040  P = 0.0001 | 0.001 | -0.038  P < 0.0001 |
| fit_ss_dayt_ave_met_value | -0.005  P = 0.6301 | 1.0 | 0.041  P < 0.0001 |
| fit_ss_total_sedentary_min | -0.017  P = 0.094 | 1.0 | -0.081  P < 0.0001 |
| fit_ss_total_ave_met | -0.0008  P = 0.937 | 1.0 | 0.056  P < 0.0001 |
| fit_ss_fitbit_sedentarymin | 0.0167  P = 0.101 | 1.0 | 0.070  P < 0.0001 |
| fit_ss_dayt_sedentary_min | -0.0049  P = 0.626 | 1.0 | -0.041  P < 0.0001 |
| fit_ss_fitbit_restingheartrate | 0.030  P = 0.003 | 0.045 | 0.119  P < 0.0001 |

| **Table 3.** Participant’s Characteristics: ADHD+ group | | |
| --- | --- | --- |
| **Description** | **M** | **SD** |
| *n = 225* |  |  |
| **Age** |  |  |
| Overall | 9.45 | 0.50 |
|  | **n** | **%** |
| **Gender** |  |  |
| Male | 143 | 63.55 |
| Female | 81 | 36.00 |
| Other | 1 | 0.44 |
| **Race** |  |  |
| White | 194 | 86.22 |
| African American | 22 | 9.77 |
| Chinese | 1 | 0.44 |
| American Indian | 1 | 0.44 |
| Asian Indian | 0 | 0.00 |
| Other | 7 | 3.11 |
| **Ethnicity** |  |  |
| Not Hispanic | 181 | 80.44 |
| Hispanic | 37 | 16.44 |
| Refused to answer | 2 | 0.88 |
| Don't know | 5 | 2.22 |
| **Parent’s Education** |  |  |
| Bachelor’s degree | 70 | 31.11 |
| Master’s degree | 56 | 24.88 |
| Some college | 40 | 17.77 |
| Associate degree | 30 | 13.33 |
| High school | 16 | 7.11 |
| Professional degree | 7 | 3.11 |
| Doctoral degree | 4 | 1.77 |
| GED or equivalent Diploma | 2 | 0.88 |
| Refused to answer | 0 | 0.00 |
| **Parent’s Income Level** |  |  |
| $100,000 to $199,999 | 72 | 32.00 |
| $50,000 to $99,999 | 67 | 29.77 |
| $200,000 and greater | 27 | 12.00 |
| $25,000 to $49,999 | 23 | 10.22 |
| Less than $25,000 | 20 | 8.88 |
| Refuse to answer | 9 | 4.00 |
| Don't know | 7 | 3.11 |
| Notes: M = Mean, SD = Standard Deviation  n is the number of participants | | |

| **Table 4.** Participant’s Characteristics: ADHD- (control) group | | |
| --- | --- | --- |
| **Description** | **M** | **SD** |
| *n = 225* |  |  |
| **Age** |  |  |
| Overall | 9.45 | 0.50 |
|  | **n** | **%** |
| **Gender** |  |  |
| Male | 114 | 50.66 |
| Female | 110 | 48.88 |
| Other | 1 | 0.44 |
| **Race** |  |  |
| White | 174 | 77.33 |
| African American | 38 | 16.88 |
| Chinese | 2 | 0.88 |
| American Indian | 1 | 0.44 |
| Asian Indian | 1 | 0.44 |
| Other | 9 | 4.00 |
| **Ethnicity** |  |  |
| Not Hispanic | 188 | 83.11 |
| Hispanic | 36 | 16.00 |
| Refused to answer | 0 | 0.00 |
| Don't know | 2 | 0.88 |
| **Parent’s Education** |  |  |
| Bachelor’s degree | 60 | 28.00 |
| Master’s degree | 58 | 25.77 |
| Some college | 30 | 13.33 |
| Associate degree | 27 | 12.00 |
| High school | 23 | 10.22 |
| Professional degree | 11 | 4.88 |
| Doctoral degree | 7 | 3.11 |
| GED or equivalent Diploma | 4 | 1.77 |
| Refused to answer | 2 | 0.88 |
| **Parent’s Income Level** |  |  |
| $100,000 to $199,999 | 67 | 29.77 |
| $50,000 to $99,999 | 54 | 24.00 |
| $200,000 and greater | 31 | 13.77 |
| $25,000 to $49,999 | 30 | 13.33 |
| Less than $25,000 | 24 | 10.66 |
| Refuse to answer | 13 | 5.77 |
| Don't know | 6 | 2.66 |
| Notes: M = Mean, SD = Standard Deviation  n is the number of participants | | |

| **Table 5.** Multivariable logistic regression model summary (with PCA) | | | | |
| --- | --- | --- | --- | --- |
| Variable | Coef | Std Err | Z-Score | P> \|Z\| |
| Intercept | -0.057877 | 0.020271 | -2.855 | 0.004302 |
| time | -0.10517 | 0.021545 | -4.881 | 1.05E-06 |
| fit_ss_wk_average_met_value | 0.147425 | 0.038766 | 3.803 | 0.000143 |
| fit_ss_wk_avg_sedentary_min | 0.050069 | 0.036824 | 1.36 | 0.173924 |
| fit_ss_fitbit_sedentarymin | 0.221149 | 0.030576 | 7.233 | 4.73E-13 |
| fit_ss_dayt_sedentary_min | -0.006154 | 0.029397 | -0.209 | 0.834192 |
| PC1 | -0.10748 | 0.018056 | -5.953 | 2.64E-09 |
| PC2 | 0.15234 | 0.020454 | 7.448 | 9.47E-14 |
| PC3 | -0.175193 | 0.097768 | -1.792 | 0.073145 |
| PC4 | 0.270596 | 0.144771 | 1.869 | 0.061604 |

| **Table 6.** Mixed-effect regression model summary | | | | |
| --- | --- | --- | --- | --- |
| Variable | Coef | Std Err | Z-Score | P > \|Z\| |
| Intercept | -14.732948 | 6.83E-01 | -21.563 | <2e-16 |
| time | -0.117312 | 0.417765 | -0.281 | 0.779 |
| fit_ss_total_sedentary_min | -0.205424 | 1.026611 | -0.2 | 0.841 |
| fit_ss_fitbit_sedentarymin | 0.075272 | 6.36E-01 | 0.118 | 0.906 |
| fit_ss_dayt_sedentary_min | 0.121566 | 0.864119 | 0.141 | 0.888 |
| fit_ss_wk_avg_sedentary_min | -0.00302 | 6.60E-01 | -0.005 | 0.996 |
| fit_ss_fitbit_rest_hr | 0.24255 | 1.56E+00 | 0.155 | 0.877 |
| fit_ss_fitbit_restingheartrate | -0.006007 | 1.537678 | -0.004 | 0.997 |
| fit_ss_wk_average_met_value | 0.05031 | 0.678025 | 0.074 | 0.941 |
| fit_ss_dayt_ave_met_value | 0.155451 | 1.305694 | 0.119 | 0.905 |
| fit_ss_total_ave_met | -0.137013 | 1.241879 | -0.11 | 0.912 |

Data processing and merging:

In our study, we utilized ABCD data release 5.0 to accurately curate our study cohorts, stratifying participants into distinct groups—those with ADHD (ADHD+) and a control group without ADHD (ADHD-). The identification of ADHD+ subjects was conducted through a comprehensive analysis of variables related to the present, past, full remission, and partial remission status of ADHD, namely 'ksads_14_856_p', 'ksads_14_853_p', 'ksads_14_854_p', 'ksads2_14_809_p', 'ksads2_14_810_p', 'ksads2_14_813_p', 'ksads_14_855_p', 'ksads2_14_811_p', and 'ksads2_14_812_p'. Our inclusion and exclusion criteria were applied to refine both the ADHD+ and ADHD- cohorts, ensuring a robust and representative selection.

In addition to the clinical ADHD data, we delved into diverse demographic factors outlined in Supplementary Material: Table 1. For the purpose of racial categorization, specific variables were consolidated and mapped into distinct categories such as White, African American, American Indian, Asian Indian, Chinese, and Other. Similar mapping procedures were executed for parental education and income levels. The resulting demographic information facilitated a stratified sampling approach to harmonize the larger ADHD- group with the ADHD+ group, after integration with Fitbit data.

The mapping schemes included detailed categorizations:

- Race mapping: {"White": 1, "African American": 2, "American Indian": 3, "Asian Indian": 4, "Chinese": 5, "Other": 6}
- Marital status mapping: {"Married": 1, "Widowed": 2, "Divorced": 3, "Separated": 4, "Never married": 5, "Living with partner": 6, "Refused to answer": 7}
- Income mapping: {"Less than $25,000": 1, "$25,000 to $49,999": 2, "$50,000 to $99,999": 3, "$100,000 to $199,999": 4, "$200,000 and greater": 5, "Don't know": 6, "Refuse to answer": 7}
- Gender mapping: {"Male": 1, "Female": 2, "Other": 3}
- Education mapping: {"Never attended": 0, "High school": 1, "GED or equivalent Diploma": 2, "Some college": 3, "Associate degree": 4, "Bachelor degree": 5, "Master degree": 6, "Professional degree": 7, "Doctoral degree": 8, "Refused to answer": 9}

Then for the Fitbit measurements, we selected specific variables, as detailed in Table 1, aligning with the ABCD data. Fitbit data collected at baseline, 2-year follow-up, and 4-year follow-up intervals were used for our analysis. The Fitbit Charge 2 model was incorporated as wrist-worn devices in the ABCD study to examine patterns of different physical activities and sleep over time with consent taken from the parents. Participants wore the Fitbit consistently for a period over 21 days, with the exception of times during bathing or water activities. The data went a comprehensive preprocessing pipeline, involving the conversion of string formats to numeric representations, the handling of date variables, merging data from different phases of data collections using 'src_subject_id', 'eventname', and 'fit_ss_day_wkno', and the removal of duplicate or null records. To address missing values, we implemented imputation techniques, replacing missing values with the mean of the respective variable for each subject, ensuring the integrity and completeness of our dataset.

Top of Form

Bottom of Form

The Fitbit data was then organized in a long format, where each row represented an independent observation, capturing daily measures for each unique participant over a 21-day period. Weekly averages were appropriately merged with daily observations based on week number and day of the week.

With Fitbit data collected at baseline, the 2-year follow-up, and the 4-year follow-up, we identified 5 unique participants at baseline, 210 at the 2-year follow-up, and 83 at the 4-year follow-up, all diagnosed with ADHD+. Overlapping participants were noted between different phases. We included a total of 225 unique ADHD+ individuals across all three Fitbit data collection waves. Some of the participants had more than 21 days of data because they were participated in more than one phase of the Fitbit data collection stages. We kept additional days because this may capture a more comprehensive and nuanced perspective of individual behaviors and trends over time.

Subsequently, we stratified and selected a cohort of 225 ADHD- subjects, matching their characteristics with those of ADHD+ subjects based on demographics. After merging ADHD diagnosis, demographics, and relevant Fitbit measurements based on 'src_subject_id', we randomized observations to create a final dataset comprising 10,045 Fitbit data records for 450 participants over 21 days across various variables.
